# The discrete update epidemics: demography, vaccination and transmission with a tensorized update approach

**DOI:** 10.1101/2022.12.10.22283299

**Authors:** Xiaohao Guo, Zeyu Zhao, Shiting Yang, Yichao Guo, Tianmu Chen

**Affiliations:** School of Public Health, State Key Laboratory of Molecular Vaccinology and Molecular Diagnostics/Xiamen University/ Xiamen City/ 361102, Fujian Province/ People’s Republic of China

## Abstract

An ordinary differential dynamical model is developed to describe the transmission of infectious diseases, considering heterogeneities of region, age, vaccination, and immigration and real-time vaccination simultaneously using a tenderized formulation. Numerical experiments are performed in Xiamen city, China, with the whole population partitioned into 6 regions × 4 age groups × 4 vaccination status groups, showing the numerical stability of the developed model. The heterogeneity consideration makes our model adequate to evaluate specific interventions accurately within specific sub-populations and carry them out.

**Author summary:** Not applicable.

## Introduction

In infectious disease modeling, the ordinary differential equations (ODEs) model focus on the state variables defined to illustrate the time-varying status of the population. Classic Susceptible-Infectious-Removed (SIR) model, Susceptible-Exposed-Infectious-Removed (SEIR) model, and Susceptible-Exposed-Symptomatic/Asymptomatic Infectious-Removed (SEIAR) model saves and updates the variable of population size of each stage of the natural history of the disease. The multi-group dynamical model performs further partitions each variable of the natural history of the SIR, SEIR, SEIAR, or other frameworks by age groups cite(models with only age groups), vaccination status groups cite(models with only vaccination status groups) or regions respectively, however, none of existing study considers those factors simultaneously with a generalized extensible formularization.

This study considers a dynamical system with age, vaccination and region factors that involving a series updates (e.g. demographic update including birth, death and growth; vaccine update including vaccinate and vaccine efficacy decay; transmission update) Most updates are modeled as linear, with the only non-linear update of transmission and vaccination. A discrete version of difference equations are firstly developed, then rewrited as differential equations. The systems of equations are formularized using tensor operations, which makes the extension of state variables to higher dimensions be possible. Numerical experiments of model simulations support the robustness of our model. A real-world scenario is chosen for model application.

## Materials and methods

The ODEs depict the relation between states and the rate of state changes and have always been solved using numerical methods for simulation or calibration.

When a ODE model is built for a certain problem, the following model simulating and model fitting process un-avoidably requires numerical methods which discretize the continuous model into a discrete one that could be solved by a computer program. In this sense, one can always build a discrete model using matrix and tensor operations at the first step, skipping the process of discretizing.

For this consideration, we will introduce the discrete version of the forward Euler method formulation naturally with detail modeling assumptions, then rewrite them as its continuous form.

### Basic tensor concepts

Assumed the basic tensor concepts of fiber and slice are familiar to readers.

**Table 1.**
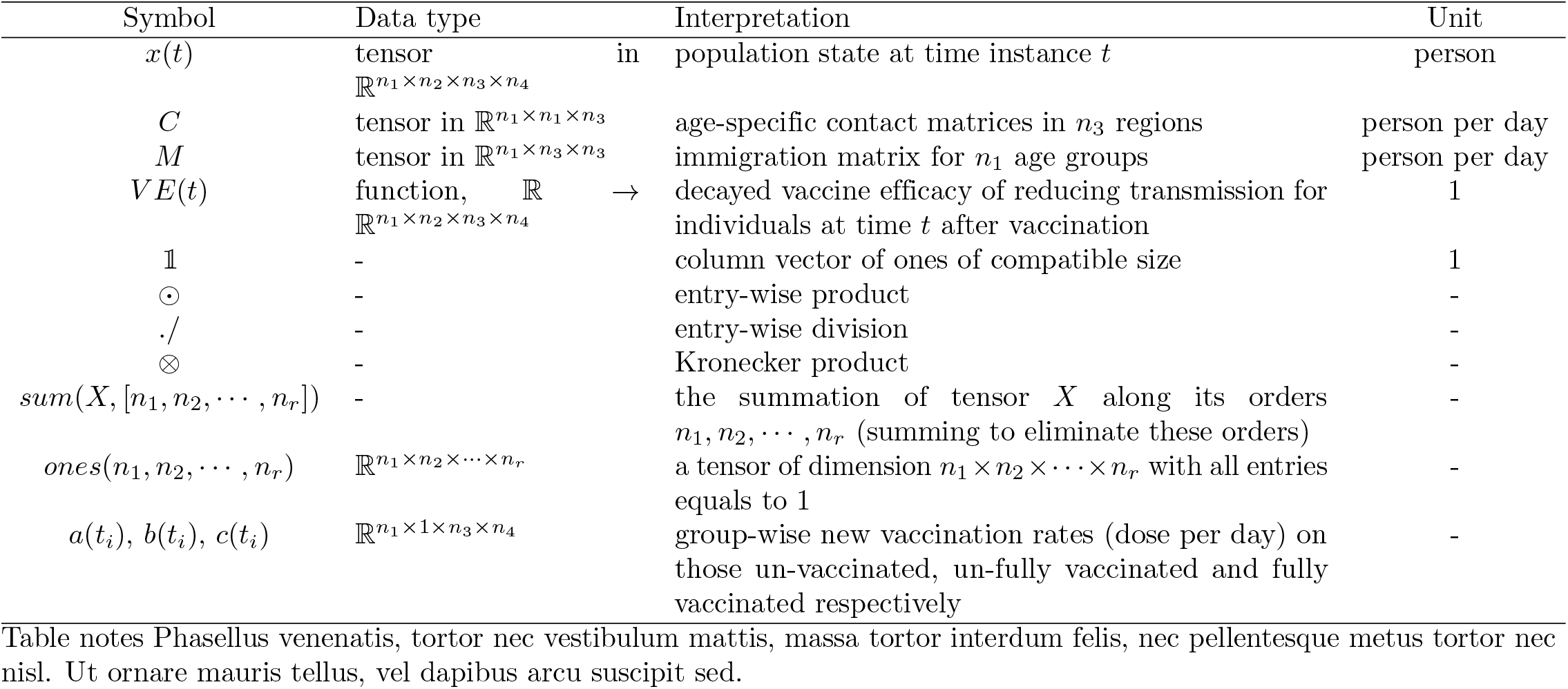
Symbols of variables and parameters

### State variable

Epidemiology studies the distribution and determinants of disease among population, where the time-varying distribution of disease could be expressed in terms of state variables of population.

Let the set of whole population 𝒩 be partitioned into the union of non-intersection sub-sets of *n*_1_ age groups, *n*_2_ vaccination status groups, *n*_3_ regions and *n*_4_ stages in natural history of disease:

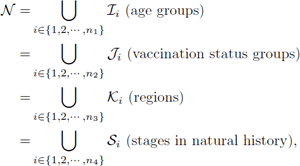

where the right-hand side is non-intersected unions of sub-sets.

We define the state variable 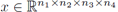, whose entry *x*_*ijks*_ denote the number of elements (individuals) in subset ℐ_*i*_ ∩ 𝒥_*j*_ ∩ 𝒦_*k*_ ∩ 𝒮_*s*_. The state variable x depicts disease distribution among the population and shall be treated as a time-dependent variable tensor.

Let [*t*_1_, *t*_2_, …, *t*_*m*_] be a discrete uniformly placed grids of time instances with *t*_*i*+1_ − *t*_*i*_ = Δ*t*, ∀*i*. The deterministic or stochastic recursion of updating *x*(*t*_*i*_) to *x*(*t*_*i*+1_) uniquely determines the dynamic process and is the key we shall model.

### Demography Operators

#### Birth and death

Suppose the group-wise natural birth rate 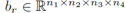 and death rate 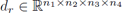 are available, then the update of birth and death is obtained:

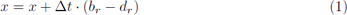

#### Growth

Growth occurs only between age groups, therefore, we shall consider for each tensor fiber 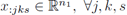, ∀*j, k, s*. Assumed the i-th age group spans over g_i_ years, e.g. the i-th group includes people aged [*g*_1_ + *g*_2_ + + *g*_*i*−1_, *g*_1_ + *g*_2_ + + *g*_*i*_), a left-close-right-open interval where g_0_ is set to be 0. Then, averagely 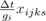 individuals will grown to the next age group after time interval Δ*t*. Such update can be described by ordinary matrix-vector produces on each tensor fiber:

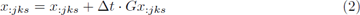

where

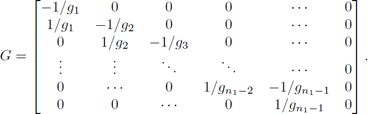

#### Immigration

The youngest and the oldest usually lesser migratable than teenagers and middle-age adults. We consider such age-specific heterogeneity in immigration. Let 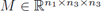 be a 3-order tensor, with its *i*-th tensor slice *M*_*i*::_ represent a matrix of immigration between n_3_ regions specifically for age group *i*. The jk-th entry of *M*_*i*::_ represent daily immigration rate of age group *i* (person per day) from region *j* to region *k*. The diagonal entries of *M*_*i*::_ are set as zeros.

Immigratiion occurs only between regions, and we shall consider for each tensor fiber *x*_*ij*:*s*_.

The proportion of *x*_*ij*:*s*_ in age group *i* of corresponding regions are given by:

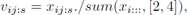

where we use the MATLAB language for convenience, i.e. sum(*X*, [*n*_1_, *n*_2_, …, *n*_*r*_]) denote summing up the *n*_1_, *n*_2_,, *n*_*r*_-th orders of tensor *X*.

For fixed age group *i*, vaccination status group *j* and natural history *s*, the immigration update is obtained:

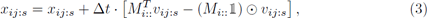

where 𝟙 is a column vector of compatible size with all entries equals 1; ⊙ represent the entry-wise product; the second term denotes immigration into regions, and shall be viewed as weighted summation of columns of *M*_*i*::_; the third term denotes immigration out regions.

In practice, 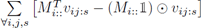 is nearly balanced to zero vector, as the immigration in and out are nearly balanced for all regions.

### Transmission Operator

We use a frequently used Susceptible (V) - Exposed (E) - Pre-symptomatic (F) - Asymptomatic (A) - Symptomatic (I) - Removed (R) framework for SARS-CoV-2 to illustrate the discrete update of transmission and stage evolution. The update is easy to degenerate to simpler SIR, SEIR, SEIAR and easy to extend to more complicated frameworks.

We arrange the *n*_4_ = 6 stages in order [V, E, F, I, A, R], i.e. *x*_:::1_ denote those susceptible, and *x*_:::6_ denote those removed.

Transmission is described by the flow rate from susceptible (S) to exposed (E). We model the force of infection 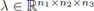 as the transmission rate coefficients acted on all susceptible subgroups *x*_:::1_.

Consider the tensor fiber *x*_:*jk*1_ of susceptible individuals with fixed vaccination status *j* and region *k*, whose *i*-th component *x*_*ijk*1_ denotes the *i*-th age group.

Assumed the age-specific contact matrices (the *ij* entry denote the daily average number of contact in age group *j* produced by one individual in age group *i*) of all *n*_3_ regions are obtained, and we save them in a tensor 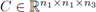. Then, for each individual in different age groups in region *k* is expected to contact the following number of symptomatic infectious individuals over a unit time span:

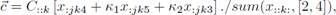

where *x*_:*jk*4_, *x*_:*jk*5_ and *x*_:*jk*3_ denote those of stage I, A, F respectively; *κ*_1_ and *κ*_2_ are relatively transmissibilities of A and F versus I (between 0 and 1); ./ denote the entry-wise division; note that the divisor is constant during updates.

Assumed each un-vaccinated susceptible individual are independently infected with probability *q* via a single contact with infectious *I*. Then, the update of population is given by:

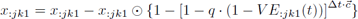

where *VE*_:*jk*1_(*t*) is the time-varying population averaging vaccine efficacy at time instance *t*, which is defined and computed via new vaccination and vaccine efficacy decays in the next two sections.

In most cases, the probability *q* is small, with the number of contact 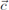 bounded, and therefore, the update can be simplified using the 1-order approximation of generalised binomial expansion. The simplified transmission update is obtained as follows:

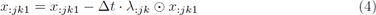

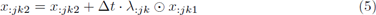

where 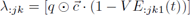, ∀*j, k* are the modeled forces of infection; the two equations represent reduction of susceptible population and increase of exposed population respectively.

### Stage Evolution Operator

In the VEFIAR framework, all parameters are potentially tensors of dimension 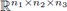, and could be considered as tensors of random variables that generate tensors of random number in each numerical update.

The update of subgroups of stage E, F, I, A, R are give by:

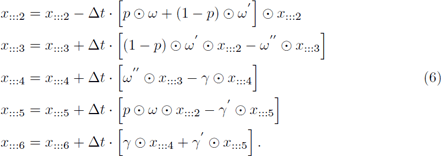

### Vaccination Operator

Assumed there are *n*_2_ = 4 vaccination status [un-vaccinated, un-fully vaccinated, fully vaccinated, booster vaccinated] under consideration. Let *a*(*t*_*i*_), *b*(*t*_*i*_), 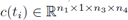 be the group-wise new vaccination rates (dose per day) on those un-vaccinated, un-fully vaccinated and fully vaccinated respectively, and are assumed as constants during time interval [*t*_*i*_, *t*_*i*+1_). People of lower vaccination status flowed in the next higher vaccination status after been vaccinated. Therefore, we define *n*_2_ = 4 tensors of unit vaccine update:

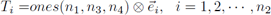

where ones(*n*_1_, *n*_3_, *n*_4_) denotes a tensor of dimension n_1_ × n_3_ × n_4_ with all entries equals to 1; ⊗ is the Kronecker product; 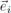 denotes the unit column vector, whose entries are zeros except 1 at the i-th entry; the orders of resulted *T*_*i*_, *i* = 1, 2,, *n*_2_ are then rearranged such that they have the same dimension as state variable 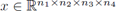.

The update of population status by newly vaccination is then obtained:

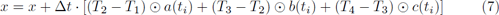

### Population Averaging Vaccine Efficacy

Although we may use empirically observed population averaging odd ratios *OR*(*t*) replacing vaccine efficacy of 1 − *VE*(*t*) in the model and simulate, here we shall consider a question of how the vaccination process and individual vaccine efficacy shape the population averaging vaccine efficacy.

Assumed we know the vaccination history, i.e., discrete *a*(*t*_*i*_), *b*(*t*_*i*_), and *c*(*t*_*i*_) for all time interval [*t*_*i*_, *t*_*i*+1_), and the empirical decay curves of individual vaccine efficacy 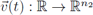 (n_2_ = 4 entries represent 4 different vaccination status).

Then, the group-wise population-averaging vaccine efficacy is obtained:

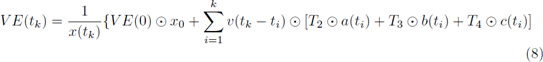

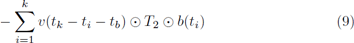

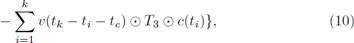

where *t*_*b*_, *t*_*c*_ denotes the average duration between the first and second dose, and between the second and third dose; the first term represents the initial vaccine efficacy among the population; the last terms are convolutions of vaccinating history and the decay of vaccination; positive terms represent the increase of population size in higher vaccination status due to vaccination; negative terms represent the decrease of population size in lower vaccination status due to vaccination (those people are vaccinated into higher vaccination status).

The modeled *VE*(*t*_*k*_) is not recursive, but involves complete history of vaccination, which brings us a great computation burden. To reduce the computational complexity in practice, we shall consider the truncation of vaccine decays. Assumed the decay curves *υ*(*t*) can be treated as constant *υ*_∞_ after a time duration of *t*_*truncated*_. Then, with the constant step size Δ*t*, we can derive the current *VE*(*t*_*k*_) using vaccination history of previous 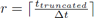 iterations, where ⌈· ⌉ denotes rounding toward positive infinity, i.e. replacing summations in Eq 10 as it starts from *i* = *r* to *k*. Such truncation provides the capability of simulating with steam data *a*(*t*_*k*_), *b*(*t*_*k*_), *c*(*t*_*k*_) of vaccination.

The immigration operator is then acted on

### Continuous version

#### Demography

##### Birth and Death

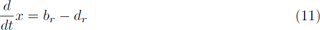

##### Growth

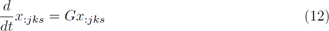

##### Immigration

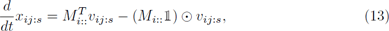

#### Transmission

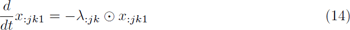

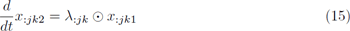

#### Stage Evolution

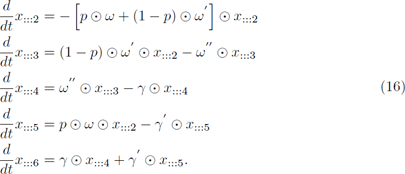

## Results

**Fig 1.**
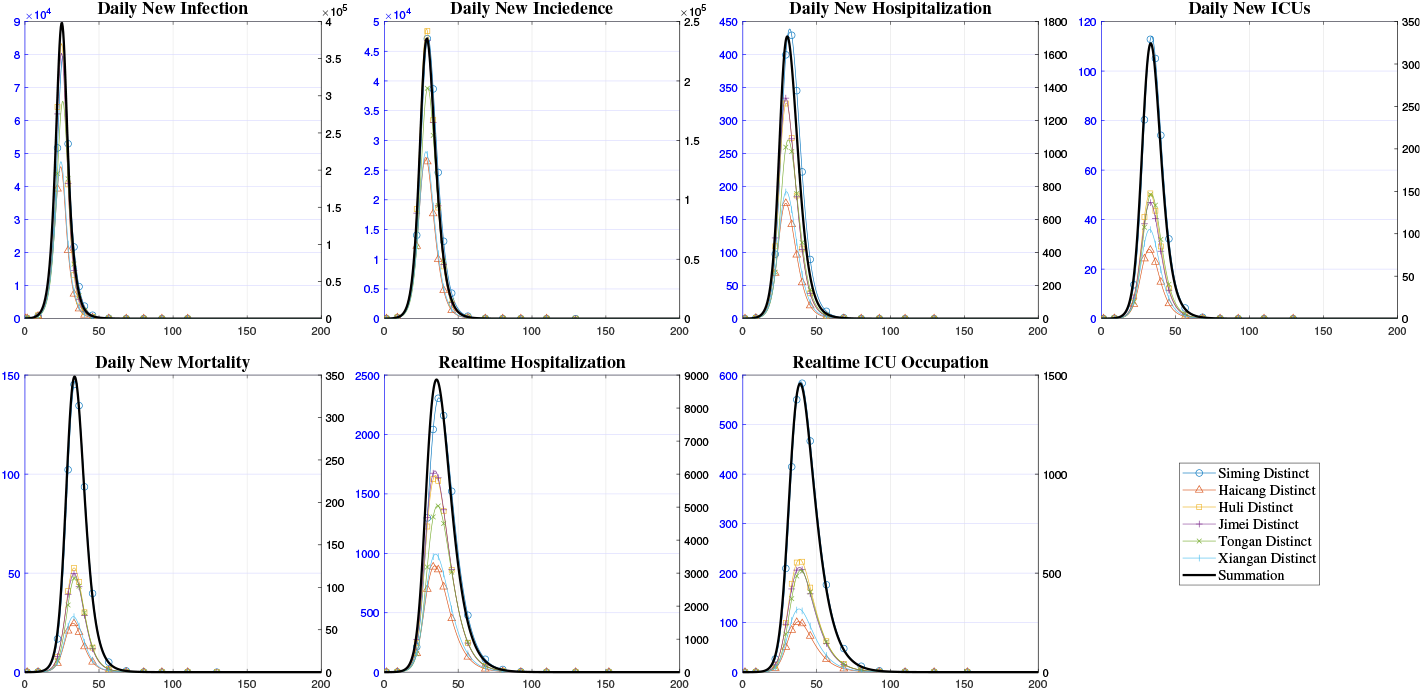
Simulated Curves By Regions. Region-wise curves are obtained by summing up dimensions of age groups and vaccination status groups.

## Discussion

Factors of disease differ significantly between age groups, vaccination groups, and regions. Using the current SARS-Cov-2 as an example, the natural history parameters of incubation, latent period and infectious period differs in vaccination status, and the regional heterogeneity of varying levels of contact frequencies and non-average intervention intensity, which have a great impact on the transmission; the risks of hospitalization, ICU ratios, and mortality concentrate on certain sub-groups of the population, presents us a series of priorities in emerging reactions.

**Fig 2.**
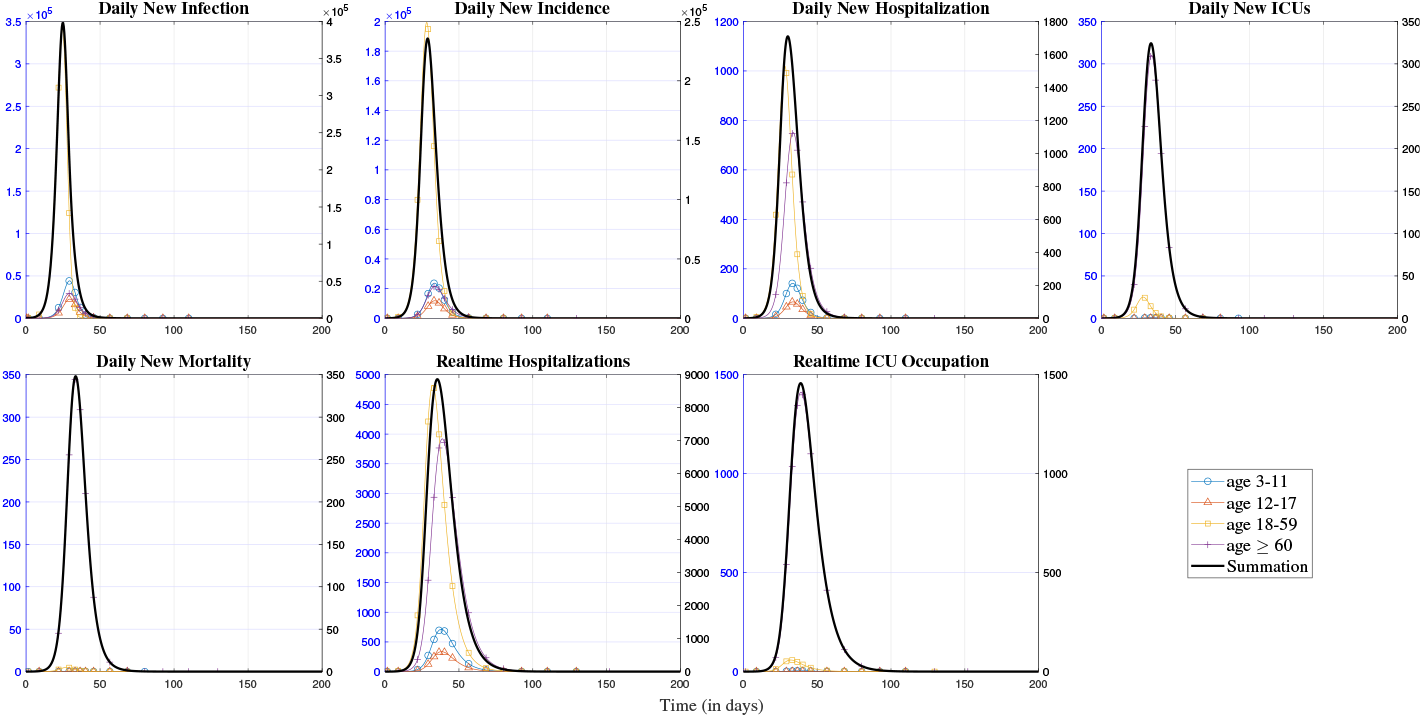
Simulated Curves By Age Groups. Age-Group-wise curves are obtained by summing up dimensions of regions and vaccination status.

## Conclusion

The developed tensorized ODE model partitions population into sub-groups of the region, age, and vaccination, considering demographic growth and immigration, and real-time vaccination progress, which could be used to evaluate a great many decision objectives. The tensorized formularization provides the capability to extend and apply our model to higher dimensions of state variables, i.e. circumstances with more sub-groups.

## Data Availability

All data produced in the present study are available upon reasonable request to the authors.

## Supporting information

## Acknowledgments

This study is supported by Self-supporting Program of Guangzhou Laboratory, Grant No. SRPG22-007.

